# Quality, Readability, and Content Deficiencies of Three Large Language Models in Bee Sting Health Information: A Comparative Evaluation with Patient Perspectives

**DOI:** 10.64898/2026.09.29.26364282

**Authors:** Renlu Xu, Lingling Yang, Xiaoling Zhang, Mula Cang, Jieru Lin, Xiaolu Jiang

**Affiliations:** People’s Hospital of Yuechi County,Guangan,638000, China; North Sichuan Medical College, Nanchong, 637000, China; Department of Emergency, Affiliated Hospital of North Sichuan Medical College, Nanchong, 6 37000, China

**Keywords:** large language models, bee sting, anaphylaxis, information quality, readability, artificial intelligence

## Abstract

**Objective:** To evaluate the information quality, reliability, readability, and content deficits of ChatGPT, DeepSeek, and Doubao in answering frequently asked questions about bee stings.

**Methods:** Twenty-five high-frequency patient questions were selected based on Google Trends, domestic health platforms, and allergy guidelines, and submitted to the three LLMs, yielding 75 responses. Two emergency physicians independently assessed quality using GQS, mDISCERN, QAMAI, EQIP, and PEMAT-U/A. Readability was evaluated using six indices (ARI, FRES, GFI, FKGL, CLI, SMOG). Content deficits were classified qualitatively. Twenty emergency department patients evaluated the comprehensibility and credibility of AI responses.

**Results:** Significant differences were found among the three models on all quality metrics (all P < 0.01). DeepSeek achieved the highest mDISCERN score (median 21.00); ChatGPT led on QAMAI (25.00) and EQIP (75.00%); Doubao ranked lowest on all quality indicators but highest on PEMAT-U (73.00%). No model met the sixth-grade reading level standard. Eleven content deficits were identified: outdated medical advice (45.45%), omissions (36.36%), and hallucinations (18.18%). Doubao received the highest patient preference (45.0%).

**Conclusion:** The three LLMs exhibited distinct strengths: DeepSeek in source reliability, ChatGPT in overall quality, and Doubao in understandability. All models had excessively high readability levels, and outdated medical advice was the predominant content deficit.

## 1. Introduction

Bee stings are among the most common acute presentations in emergency departments worldwide, with a substantial number of patients seeking care each year following hymenoptera envenomation. Bee venom contains multiple bioactive components—including melittin, phospholipase A2, and hyaluronidase—that can trigger a spectrum of clinical manifestations ranging from mild local reactions to fatal anaphylactic shock^[1]^. Approximately 5% to 7.5% of adults experience systemic allergic reactions following a bee sting during their lifetime, with a fatality rate of approximately 0.03 per million ^[1]^. For patients with a history of bee venom allergy, the risk of severe anaphylaxis upon re-sting may reach 30% to 70% ^[2]^. Accordingly, accurate and comprehensible health information about bee stings is essential for patient safety management and timely medical consultation.

In recent years, large language models (LLMs), a core technology underlying generative artificial intelligence, have been increasingly applied in health care ^[3]^. ChatGPT (OpenAI), DeepSeek, and Doubao (ByteDance) are currently the three most widely used LLMs domestically and internationally, and growing numbers of patients and members of the public rely on these platforms to obtain medical and health information ^[5]^. Existing studies have demonstrated that the quality of information produced by LLMs varies considerably across specialty domains, including orthopedics ^[6]^, obstetrics ^[7]^, dermatology ^[8]^, oncology ^[9]^, allergology ^[28]^, and emergency medicine ^[30]^, and that excessively high readability levels are a pervasive problem ^[32]^. However, systematic evaluation of LLM performance in the context of bee stings—a common emergency condition—remains lacking.

Health information about bee stings spans multiple dimensions: allergy mechanisms, first-aid management, epinephrine use, immunotherapy, and routine prevention. The content ranges from basic public education to specialized emergency care, placing high demands on LLMs with respect to information completeness, accuracy, and accessibility. Moreover, the primary audience for bee sting information comprises lay persons and patients, making readability a noteworthy concern: if AI-generated health information is excessively technical, it may fail to effectively guide patients’ self-management behavior.

This study aimed to systematically evaluate the information quality, reliability, readability, and content deficits of three mainstream LLMs—ChatGPT, DeepSeek, and Doubao—when responding to frequently asked questions about bee stings, with the goal of providing a reference for clinicians and patients and informing the optimization of LLMs in the domain of anaphylaxis first aid.

## 2. Methods

### 2.1 Study Design

This cross-sectional observational study was conducted in August 2026 in Sichuan, China. The study adhered to the Strengthening the Reporting of Observational Studies in Epidemiology (STROBE) reporting guidelines ^[10]^ and the Chatbot Assessment Reporting Tool (CHART) guidelines ^[11]^. A flowchart of the study design is presented in Figure 1.

**Figure 1.**
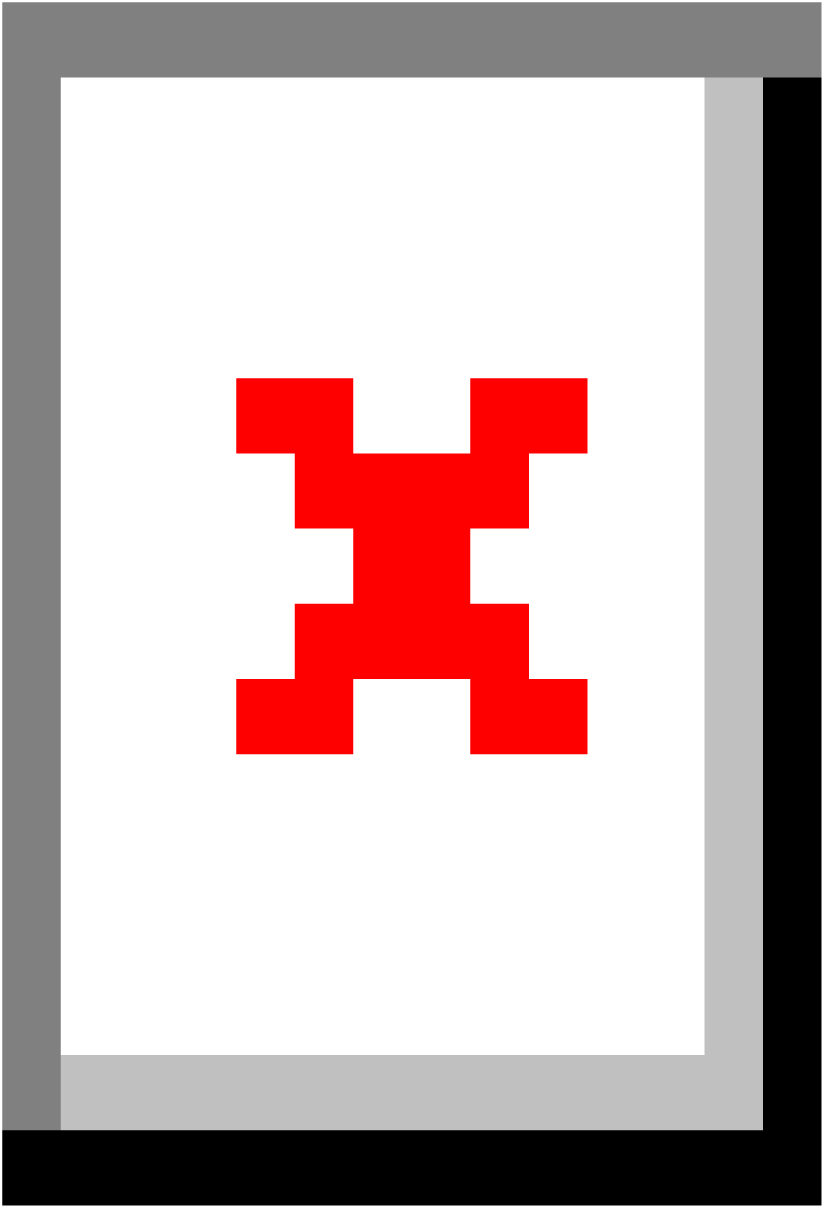
Study flowchart for the evaluation of large language model responses to bee sting–related patient questions. Twenty-five high-frequency patient questions were identified from Google Trends, domestic health platforms, and authoritative allergy guidelines, and independently submitted to ChatGPT, DeepSeek, and Doubao, yielding 75 responses. Responses were evaluated across three dimensions: quality and reliability, readability, and content deficiency. After inter-rater reliability testing (ICC), a patient survey (n = 20) was conducted, followed by statistical analysis using the Kruskal-Wallis test, one-way ANOVA, and Spearman correlation with visualization.

### 2.2 Question Selection

High-frequency questions about bee stings were identified through three channels: (1) Google Trends (2020–2026) was queried using the keywords ^“^bee sting,^” “^蜂蜇伤^”^ (bee sting injury)^, “蜜蜂蛰伤”^ (honeybee sting), and ^“^蜂毒过敏” (bee venom allergy), and the top-ranked patient questions were extracted; (2) common consultations were retrieved from major domestic health information platforms, including Baidu Health, DXY (DingXiang Doctor), and Haodf.com; and (3) core knowledge points were referenced from the Chinese Guidelines for the Diagnosis and Treatment of Allergic Reactions ^[12]^ and the World Allergy Organization (WAO) insect venom allergy guidelines ^[13]^. After deduplication, consolidation, and review by clinical experts, 25 questions were finalized.

These questions covered sting mechanisms, local reactions, systemic allergy, first-aid management, pharmacotherapy, immunotherapy, daily prevention, and rehabilitation.

### 2.3 LLM Selection and Question-Answering Procedure

Three LLMs were selected for evaluation: ChatGPT (OpenAI, version GPT-5.6), DeepSeek (version V4), and Doubao (ByteDance, version Doubao-2.1), all accessed via their official web interfaces in August 2026. Each question was entered in an independent conversation window using a standardized format to avoid contextual interference. Web search and plugin functions were disabled. Responses were recorded in their entirety as plain text, yielding a total of 75 responses (25 questions × 3 models).

### 2.4 Assessment Instruments

#### 2.4.1 Quality and Reliability Assessment

Five validated assessment instruments were employed: (1) Global Quality Scale (GQS) ^[14]^, a 5-point Likert scale evaluating the overall quality of online health information (1 = poor, 5 = excellent); (2) modified DISCERN (mDISCERN) ^[15]^, comprising 5 core items (clear aims, reliable sources, balanced and unbiased, additional sources, areas of uncertainty), each scored 1–5 for a total of 5–25; (3) Quality Analysis of Medical AI (QAMAI) ^[16]^, assessing accuracy, clarity, relevance, completeness, source referencing, and usefulness across 6 dimensions (0–5 each, total 0–30); (4) Ensuring Quality Information for Patients (EQIP) ^[17]^, comprising 20 items scored as ^“^yes^”^ (1), ^“^partially^”^ (0.5), or ^“^no^”^ (0) and expressed as a percentage; and (5) Patient Education Materials Assessment Tool for Understandability and Actionability (PEMAT-U/A) ^[18]^, evaluating understandability and actionability separately, each expressed as a percentage.

#### 2.4.2 Readability Assessment

Six widely used readability indices were calculated: Automated Readability Index (ARI) ^[19]^, Flesch Reading Ease Score (FRES) ^[20]^, Gunning Fog Index (GFI) ^[21]^, Flesch–Kincaid Grade Level (FKGL) ^[22]^, Coleman–Liau Index (CLI) ^[23]^, and Simple Measure of Gobbledygook (SMOG) ^[24]^. All readability assessments were performed using the online tool at readabilityformulas.com.

#### 2.4.3 Content Deficit Classification

Following the methodology of Zhang et al. ^[25]^, content deficits were classified into four categories: output errors, hallucinations, omissions of critical information, and outdated medical advice.

### 2.5 Assessment Procedure

Two emergency physicians from different hospitals, each with more than 5 years of clinical experience, independently assessed all responses. Prior to assessment, all responses were de-identified, randomized in sequence, and stripped of model identifiers to enable blinded evaluation. Inter-rater reliability was assessed using the intraclass correlation coefficient (ICC), with ICC < 0.50 considered poor, 0.50–0.75 moderate, 0.75–0.90 good, and > 0.90 excellent ^[26]^. Items with a scoring discrepancy of ≥ 1 point were resolved through consensus discussion.

### 2.6 Patient Survey

Five representative questions were selected from the 25 (covering basic education, safety warnings, first-aid action, medication guidance, and daily prevention), and 20 patients presenting to the emergency department were recruited between September 2 and September 14, 2026, for subjective evaluation. Inclusion criteria were age ≥ 18 years, ability to read Chinese independently, and provision of informed consent. Exclusion criteria included critical illness, cognitive impairment, and visual impairment. Informed consent was obtained from all participants prior to questionnaire completion; the consent form was presented on the first page of the electronic questionnaire, and participants could proceed to the survey items only after providing explicit agreement. Each patient read three anonymized AI responses to the same question and rated them on comprehensibility (6 items, 1–5 scale) and credibility (1 item), then selected the most preferred response.

### 2.7 Statistical Analysis

Data normality was assessed using the Shapiro–Wilk test. Normally distributed data are expressed as mean ± standard deviation (Mean ± SD), and non-normally distributed data as median (interquartile range) [M (Q₁, Q₃)]. Multi-group comparisons were performed using the Kruskal–Wallis test (non-normal distribution) or one-way analysis of variance (normal distribution), with post hoc pairwise comparisons using Bonferroni-corrected Mann–Whitney U tests or Tukey HSD tests. P value < 0.05 was considered statistically significant. All statistical analyses were conducted using R version 4.5.2.

## 3. Results

### 3.1 Inter-Rater Reliability

Inter-rater agreement across all assessment instruments ranged from moderate to excellent. QAMAI yielded the highest ICC at 0.953 (95% CI: 0.927–0.970), followed by mDISCERN at 0.917 (95% CI: 0.872–0.947), PEMAT-A at 0.824 (95% CI: 0.735–0.885), PEMAT-U at 0.731 (95% CI: 0.605–0.821), GQS at 0.725 (95% CI: 0.597–0.817), and EQIP at 0.631 (95% CI: 0.473–0.750), all reaching at least moderate agreement.

### 3.2 Quality and Reliability Comparisons

Comparisons of quality and reliability metrics across the three models are presented in Table 3 and Figures 2. For GQS, the medians for ChatGPT and DeepSeek were both 5.00, compared with 4.00 for Doubao (P < 0.01). For mDISCERN, DeepSeek performed best (median 21.00), followed by ChatGPT (18.00) and Doubao (13.00), with statistically significant differences among the three groups (P < 0.01). For QAMAI, ChatGPT achieved the highest score (median 25.00), followed by DeepSeek (24.00) and Doubao (20.00) (P < 0.01). For EQIP, the medians were 75.00% for ChatGPT, 70.00% for DeepSeek, and 62.50% for Doubao (P < 0.01).

**Figure 2.**
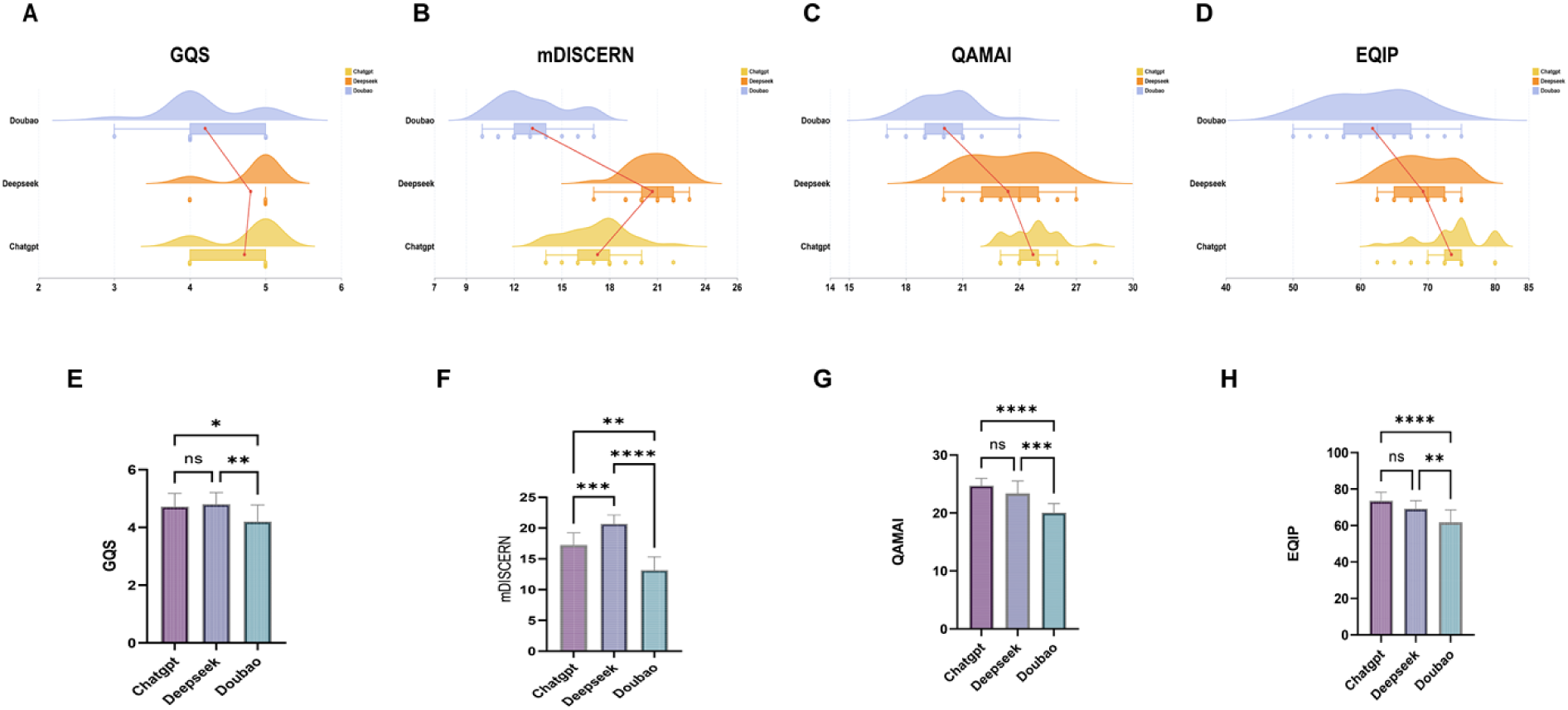
Comparison of GQS, QAMAI, mDISCERN, and EQIP scores among three large language models. A–D: Ridge plots illustrating the distribution and median trends of ChatGPT, DeepSeek, and Doubao on the GQS (A), QAMAI (B), mDISCERN (C), and EQIP (D) scales. E–H: Bar charts showing mean scores with intergroup comparisons. For GQS, no significant difference was observed between ChatGPT and DeepSeek, while both scored significantly higher than Doubao (*P < 0.05, **P < 0.01). For QAMAI, ChatGPT and DeepSeek did not differ significantly, but both were significantly higher than Doubao (***P < 0.001, ****P < 0.0001). For mDISCERN, DeepSeek achieved the highest scores, with statistically significant differences among all pairwise comparisons (**P < 0.01, ***P < 0.001, ****P < 0.0001). For EQIP, ChatGPT scored highest, with significant differences between ChatGPT and Doubao (****P < 0.0001) and between DeepSeek and Doubao (***P < 0.001), while the difference between ChatGPT and DeepSeek was not significant (ns).

**Table 1.** List of Common Patient Questions About Bee Stings Used as Exact Prompts Across All LLM Platforms.

| Question No. | Question Category | Question |
| --- | --- | --- |
| 1 | Etiology and Risk Factors | After I am stung by a bee, why does my body develop pain, swelling, itching, or an allergic reaction? |
| 2 | Etiology and Risk Factors | I previously had only local swelling after a sting. Could I develop a systemic allergic reaction this time? |
| 3 | Etiology and Risk Factors | What factors increase my risk of a severe allergic reaction or recurrence after a bee sting? |
| 4 | Clinical Manifestations and Severity | What local symptoms after a bee sting are usually considered a mild reaction? |
| 5 | Clinical Manifestations and Severity | How can I tell whether my local swelling is a “large local reaction” rather than an ordinary reaction? |
| 6 | Clinical Manifestations and Severity | What symptoms after a bee sting suggest that I may be having a severe allergic reaction? |
| 7 | Clinical Manifestations and Severity | If I am stung by many bees at the same time, why can it be dangerous even if I am not allergic? |
| 8 | Diagnosis and Assessment | What medical history and symptoms are used to determine whether I have bee venom allergy? |
| 9 | Diagnosis and Assessment | When do I need bee venom-specific IgE testing, a skin-prick test, or an intradermal test? |
| 10 | Diagnosis and Assessment | If I have an allergy test immediately after being stung and the result is negative, does that rule out bee venom allergy? |
| 11 | First Aid and Emergency Treatment | What should I do first after I am stung by a bee? |
| 12 | First Aid and Emergency Treatment | Why does the stinger need to be removed as soon as possible after a honeybee sting? |
| 13 | First Aid and Emergency Treatment | Should I scrape or pull the stinger out? |
| 14 | First Aid and Emergency Treatment | Can I wash the sting site with soap and water after a bee sting? |
| 15 | First Aid and Emergency Treatment | What over-the-counter medicines or cold-compress methods can I use for local pain, swelling, or itching after a bee sting? |
| 16 | First Aid and Emergency Treatment | Why do I need prompt medical attention if I am stung in or around the eye? |
| 17 | First Aid and Emergency Treatment | What symptoms after a bee sting mean that I must call emergency services or go to the emergency department immediately? |
| 18 | First Aid and Emergency Treatment | When should I use the first dose or a second dose of an epinephrine autoinjector? |
| 19 | First Aid and Emergency Treatment | If I cannot use an epinephrine autoinjector myself, can someone else help me use it? |
| 20 | Long-term Management and Venom Immunotherapy | After I have had a systemic allergic reaction caused by a bee sting, do I need to carry an emergency kit or epinephrine autoinjector long term? |
| 21 | Long-term Management and Venom Immunotherapy | Am I a candidate for venom immunotherapy? |
| 22 | Long-term Management and Venom Immunotherapy | How much can venom immunotherapy reduce my risk of another severe allergic reaction? |
| 23 | Long-term Management and Venom Immunotherapy | How long do I usually need venom immunotherapy, and in what situations might I need a longer course or even long-term treatment? |
| 24 | Prognosis, Prevention, and Daily Care | How long do local pain and swelling usually last after a bee sting? |
| 25 | Prognosis, Prevention, and Daily Care | What can I do in daily life to reduce my risk of being stung again and having another severe reaction? |

**Table 2.** Intraclass Correlation Coefficient Between Two Raters for Different Parameters.

| Metric | ICC | CI lower | CI upper | p |
| --- | --- | --- | --- | --- |
| GQS | 0.725 | 0.597 | 0.817 | <0.01 |
| mDISCERN | 0.917 | 0.872 | 0.947 | <0.01 |
| EQIP | 0.631 | 0.473 | 0.750 | <0.01 |
| QAMAI | 0.953 | 0.927 | 0.970 | <0.01 |
| PEMAT-U | 0.731 | 0.605 | 0.821 | <0.01 |
| PEMAT-A | 0.824 | 0.735 | 0.885 | <0.01 |

**Table 3.** Comparison of Response Quality and Reliability Across Different Large Language Models.

| Variables | Chatgpt | Deepseek | Doubao | p |
| --- | --- | --- | --- | --- |
| GQS, M ($Q_1$, $Q_3$) | 5.00 (4.00,5.00) | 5.00 (5.00,5.00) | 4.00 (4.00,5.00) | <0.01 |
| Mdiscern, M ($Q_1$, $Q_3$) | 18.00<br>(16.00,18.00) | 21.00<br>(20.00,22.00) | 13.00<br>(12.00,14.00) | <0.01 |
| QAMAI, M ($Q_1$, $Q_3$) | 25.00<br>(24.00,25.00) | 24.00<br>(22.00,25.00) | 20.00<br>(19.00,21.00) | <0.01 |
| EQIP, M ($Q_1$, $Q_3$) | 75.00<br>(72.50,75.00) | 70.00<br>(65.00,72.50) | 62.50<br>(57.50,67.50) | <0.01 |

### 3.3 Understandability and Actionability Comparisons

Comparisons of PEMAT-U and PEMAT-A scores are shown in Table 4 and Figure 3. For understandability, Doubao attained the highest PEMAT-U median (73.00%), followed by ChatGPT (71.00%) and DeepSeek (65.00%), with a statistically significant difference among the three groups (P < 0.01). For actionability, ChatGPT and DeepSeek both achieved a PEMAT-A median of 60.00%, compared with 40.00% for Doubao (P < 0.01).

**Figure 3.**
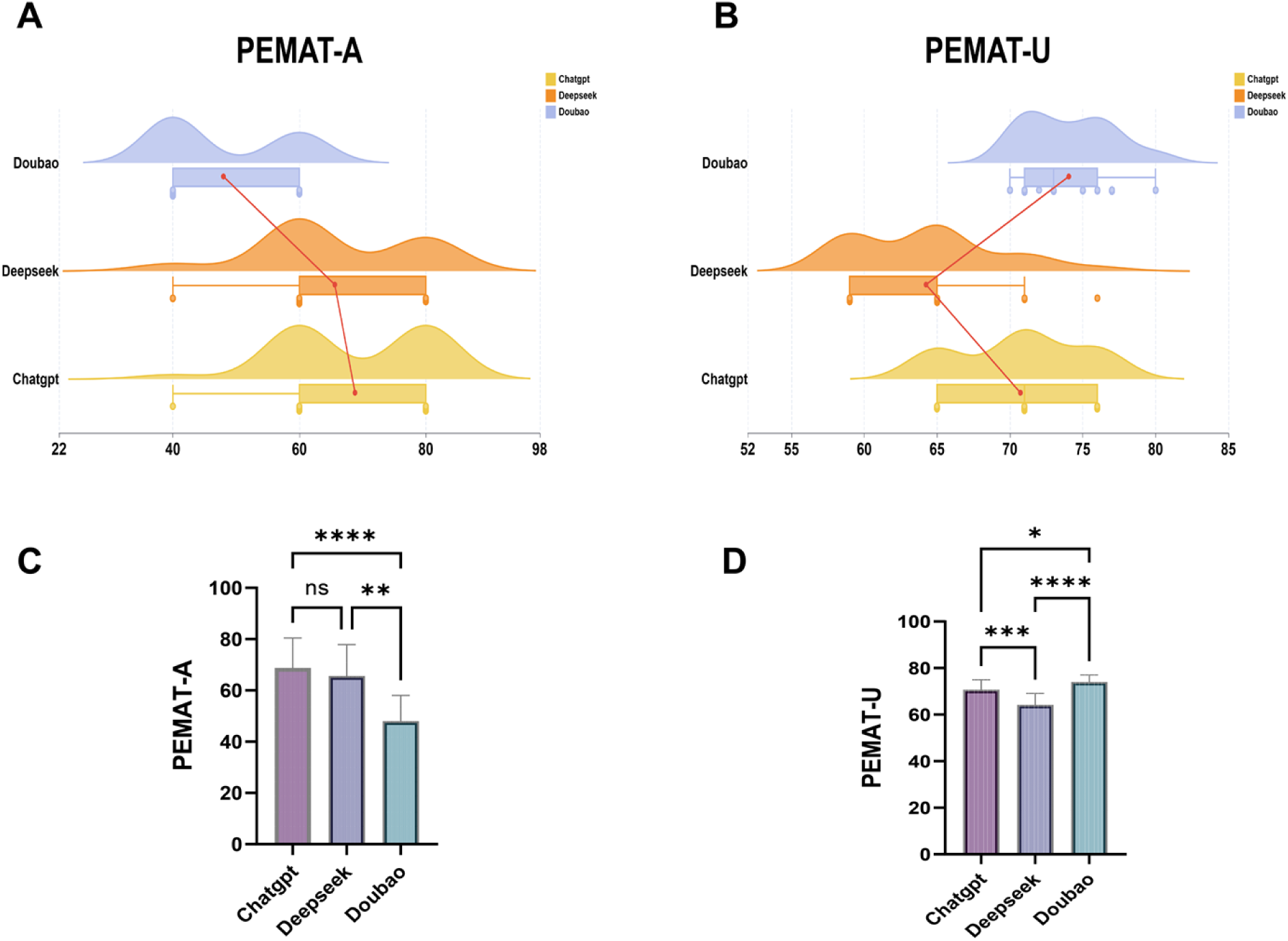
Comparison of PEMAT -A and PEMAT-U scores among three large language models. A–B: Violin plots combined with box plots illustrating the distribution of ChatGPT, DeepSeek, and Doubao on the PEMAT-A (A) and PEMAT-U (B) scales. C–D: Bar charts showing mean scores with intergroup comparisons. For PEMAT-U, Doubao achieved the highest scores, with statistically significant differences among all pairwise comparisons (*P < 0.05, ***P < 0.001, ****P < 0.0001). For PEMAT-A, no significant difference was observed between ChatGPT and DeepSeek, while both scored significantly higher than Doubao (**P < 0.01, ****P < 0.0001).

**Table 4.** Comparison of Response Understandability and Actionability Across Different Large Language Models.

| Variables | ChatGPT | Deepseek | Doubao | p |
| --- | --- | --- | --- | --- |
| PEMAT-U, M | 71.00 | 65.00 | 73.00 | <0.01 |
| (Q<sub>1</sub>, Q<sub>3</sub>) | (65.00,76.00) | (59.00,65.00) | (71.00,76.00) |  |
| PEMAT-A, M | 60.00 | 60.00 | 40.00 | <0.01 |
| (Q<sub>1</sub>, Q<sub>3</sub>) | (60.00,80.00) | (60.00,80.00) | (40.00,60.00) |  |

### 3.4 Readability Assessment

Results of the six readability indices are presented in Table 5 and Figures 4–5. ARI, FKGL, and SMOG showed statistically significant differences among the three groups (all P < 0.01), whereas FRES, GFI, and CLI did not (all P > 0.05). DeepSeek exhibited the highest ARI (13.88 ± 3.01) and FKGL (12.48 ± 2.65), indicating the greatest reading difficulty. Doubao had the lowest SMOG (9.46 ± 2.61), reflecting relatively better readability. The FKGL for all models exceeded grade 10, falling well short of the sixth-grade reading level recommended by the American Medical Association.

**Figure 4.**
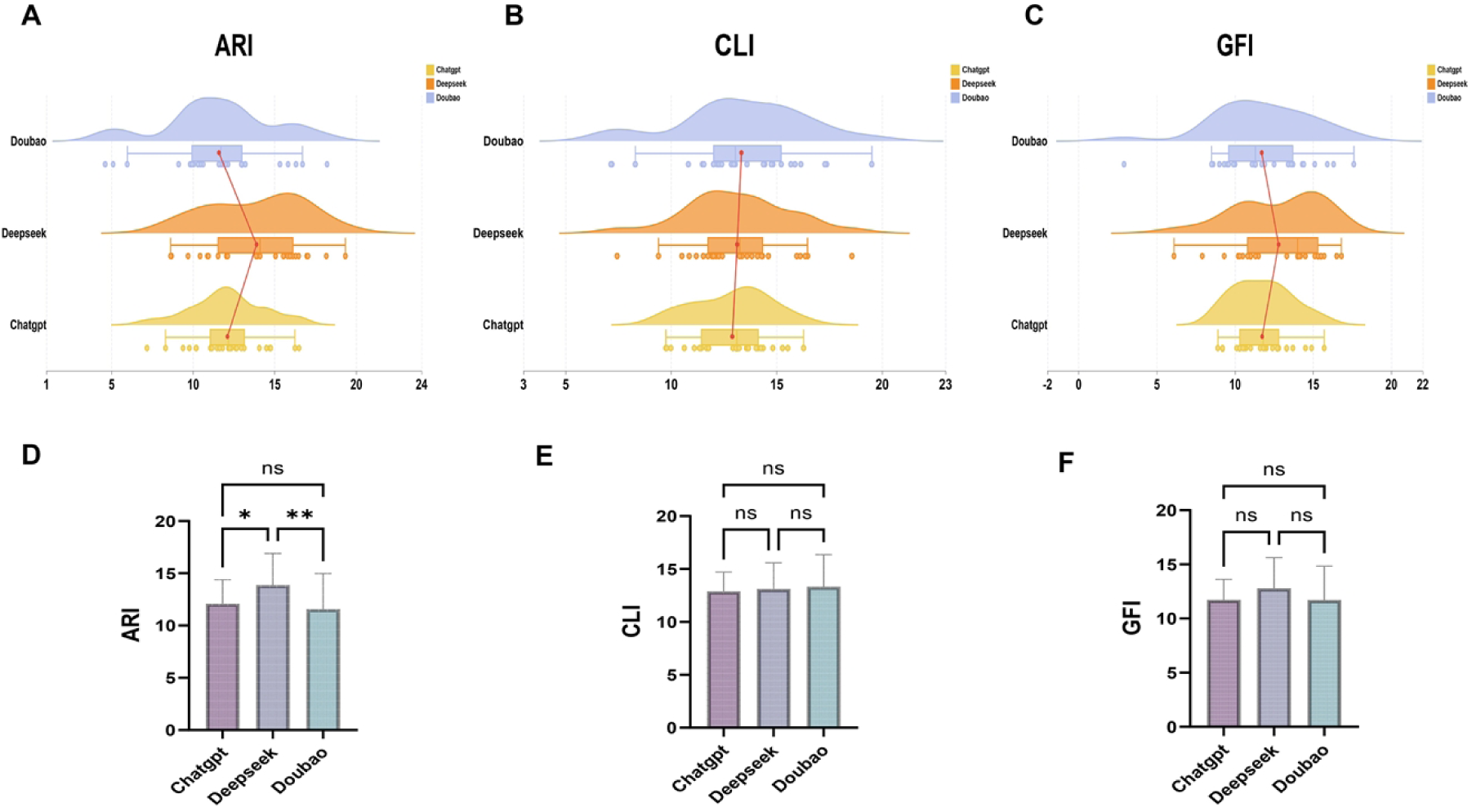
Comparison of ARI, CLI, and GFI readability indices among three large language models. A–C: Violin plots combined with box plots illustrating the distribution of ChatGPT, DeepSeek, and Doubao on the ARI (A), CLI (B), and GFI (C) indices. D–F: Bar charts showing mean scores with intergroup comparisons. For ARI, DeepSeek scored significantly higher than ChatGPT (*P < 0.05) and Doubao (**P < 0.01), with no significant difference between ChatGPT and Doubao. For CLI and GFI, no significant differences were observed among any pairwise comparisons (ns).

**Figure 5.**
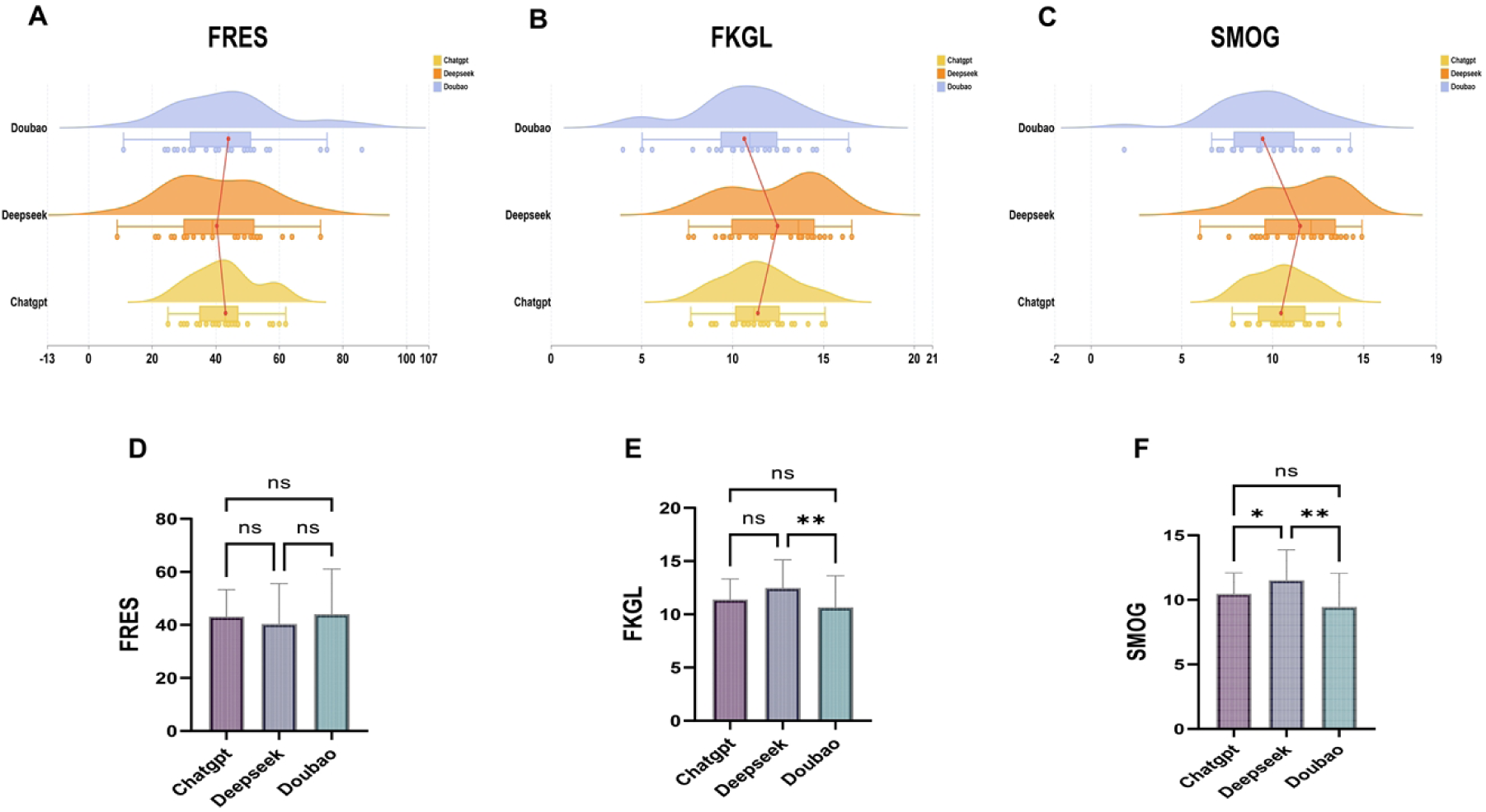
Comparison of FRES, FKGL, and SMOG readability indices among three large language models. A–C: Violin plots combined with box plots and scatter points illustrating the distribution of ChatGPT, DeepSeek, and Doubao on the FRES (A), FKGL (B), and SMOG (C) indices. D–F: Bar charts showing mean scores with intergroup comparisons. For FRES, no significant differences were observed among any pairwise comparisons (ns). For FKGL, both ChatGPT and DeepSeek scored significantly higher than Doubao (**P < 0.01), with no significant difference between ChatGPT and DeepSeek. For SMOG, statistically significant differences were found across all pairwise comparisons (*P < 0.05, **P < 0.01).

**Table 5.**
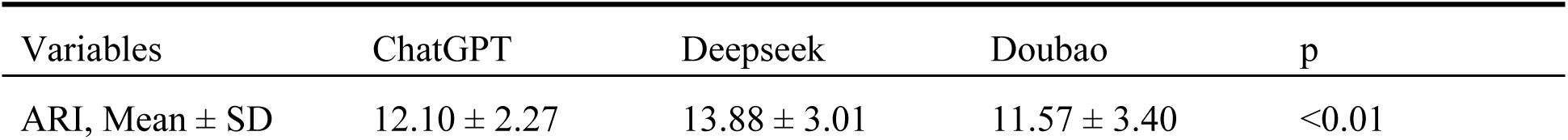
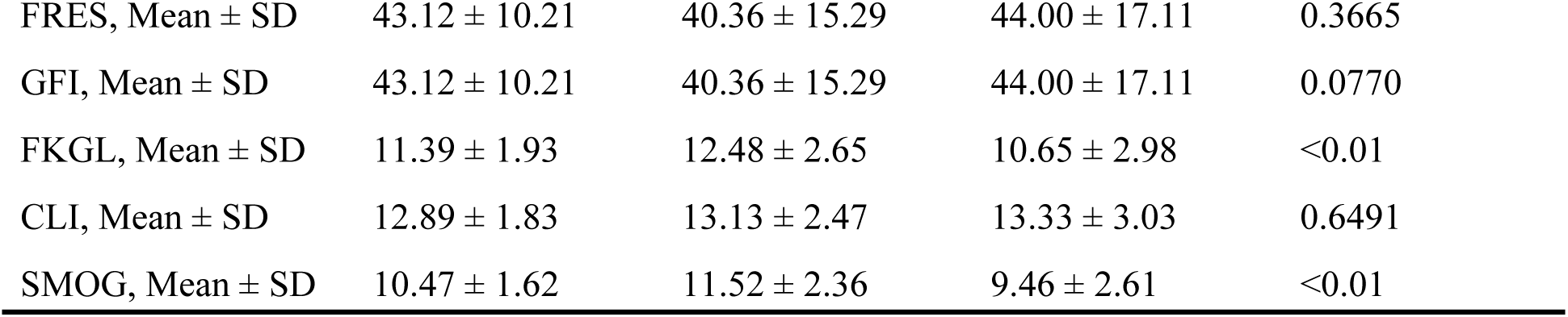
Comparison of Response Readability Across Different Large Language Models.

| Variables | ChatGPT | Deepseek | Doubao | p |
| --- | --- | --- | --- | --- |
| ARI, Mean ± SD | 12.10 ± 2.27 | 13.88 ± 3.01 | 11.57 ± 3.40 | <0.01 |
| FRES, Mean ± SD | 43.12 ± 10.21 | 40.36 ± 15.29 | 44.00 ± 17.11 | 0.3665 |
| GFI, Mean ± SD | 43.12 ± 10.21 | 40.36 ± 15.29 | 44.00 ± 17.11 | 0.0770 |
| FKGL, Mean ± SD | 11.39 ± 1.93 | 12.48 ± 2.65 | 10.65 ± 2.98 | <0.01 |
| CLI, Mean ± SD | 12.89 ± 1.83 | 13.13 ± 2.47 | 13.33 ± 3.03 | 0.6491 |
| SMOG, Mean ± SD | 10.47 ± 1.62 | 11.52 ± 2.36 | 9.46 ± 2.61 | <0.01 |

### 3.5 Content Deficit Analysis

The classification of content deficits across the 75 responses is shown in Table 6 and Figure 6. Among the 75 responses, 11 content deficits were identified. Outdated medical advice was the most common deficit type, accounting for 45.45% (5 instances) and involving management practices no longer recommended. Omissions of critical information constituted 36.36% (4 instances), with the missing information potentially affecting patients’ decisions to seek care. Hallucinations accounted for 18.18% (2 instances), manifesting as fabricated content lacking an evidence base. No output errors were detected.

**Figure 6.**
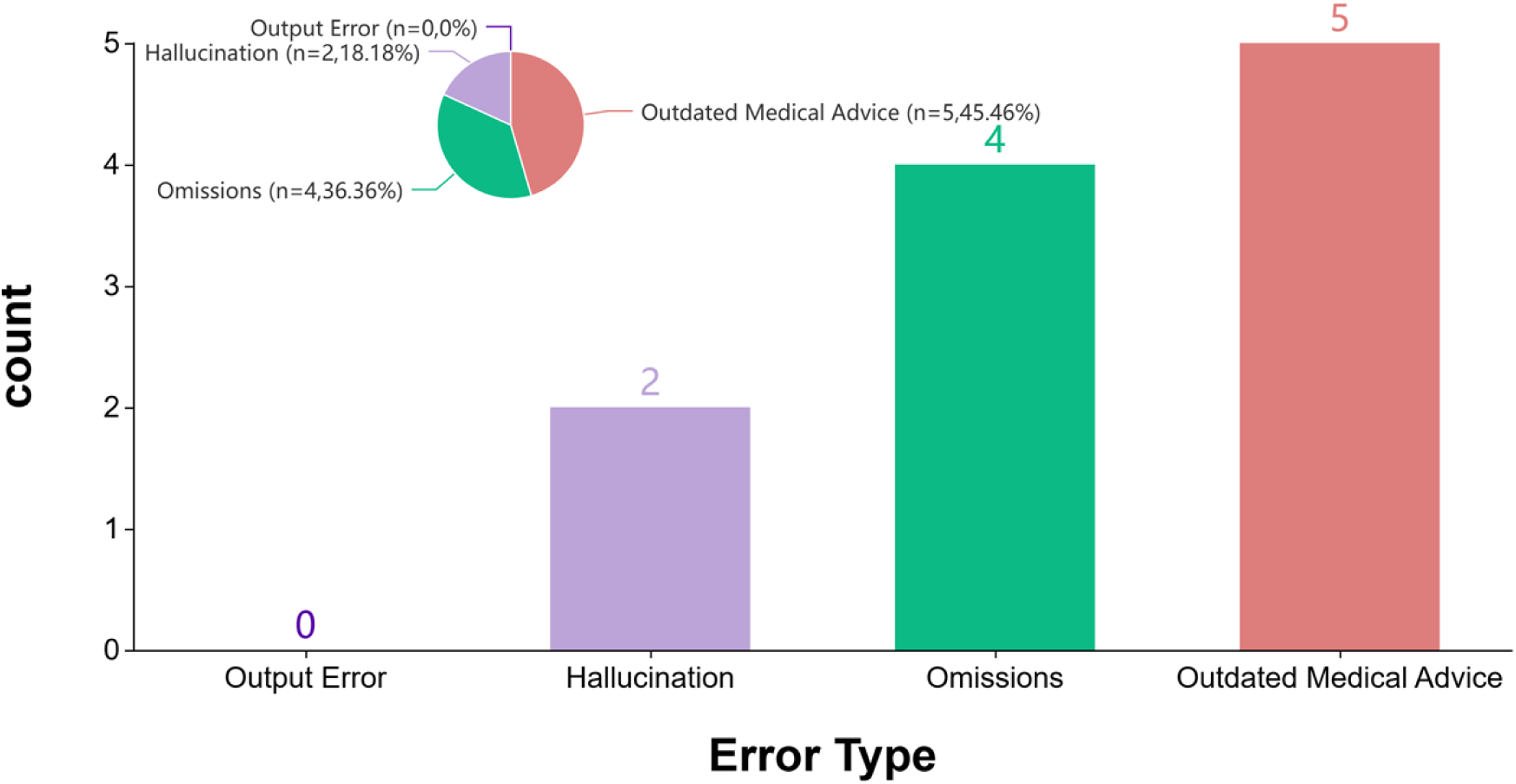
Distribution of content deficiency types in AI-generated responses. Bar chart displaying the frequency of four content deficiency categories, with a pie chart showing their proportional composition. Among 75 AI responses, a total of 11 content deficiencies were identified: 5 cases of outdated medical advice (45.46%), 4 cases of omissions (36.36%), 2 cases of hallucinations (18.18%), and 0 cases of output errors (0.00%).

**Figure 7.**
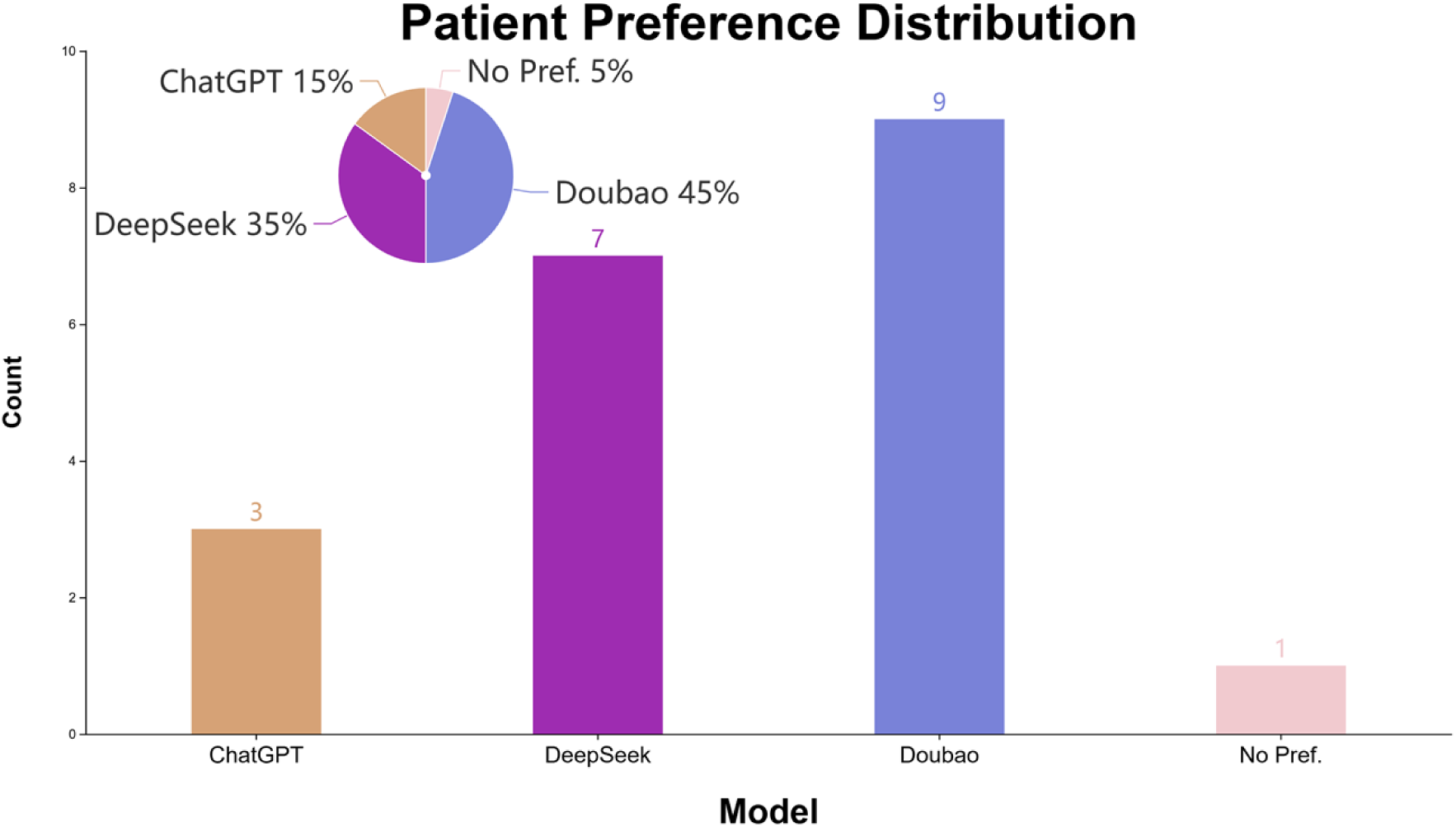
Distribution of patient preferences for responses from three large language models. Bar chart and pie chart displaying the preference distribution among 20 patients. Nine patients (45%) preferred Doubao, 7 (35%) preferred DeepSeek, 3 (15%) preferred ChatGPT, and 1 (5%) expressed no preference. Comprehensibility scores (6 items, 1–5 scale) for the three AI responses were as follows: ChatGPT, 3.62 ± 0.38; DeepSeek, 3.74 ± 0.28; and Doubao, 3.95 ± 0.27. The Friedman test indicated a statistically significant difference among the three groups (χ² = 8.351, *P* = 0.015). Post hoc pairwise comparisons showed that Doubao significantly outperformed both ChatGPT (Pcorrected = 0.041) and DeepSeek (Pcorrected = 0.041), while ChatGPT and DeepSeek did not differ significantly (Pcorrected = 1.000). Credibility scores were 3.35 ± 1.09 for ChatGPT, 3.90 ± 0.55 for DeepSeek, and 4.15 ± 0.59 for Doubao, with a statistically significant difference among the three groups (χ² = 6.241, *P* = 0.044); Doubao outperformed ChatGPT (Pcorrected = 0.028).

**Figure 8.**
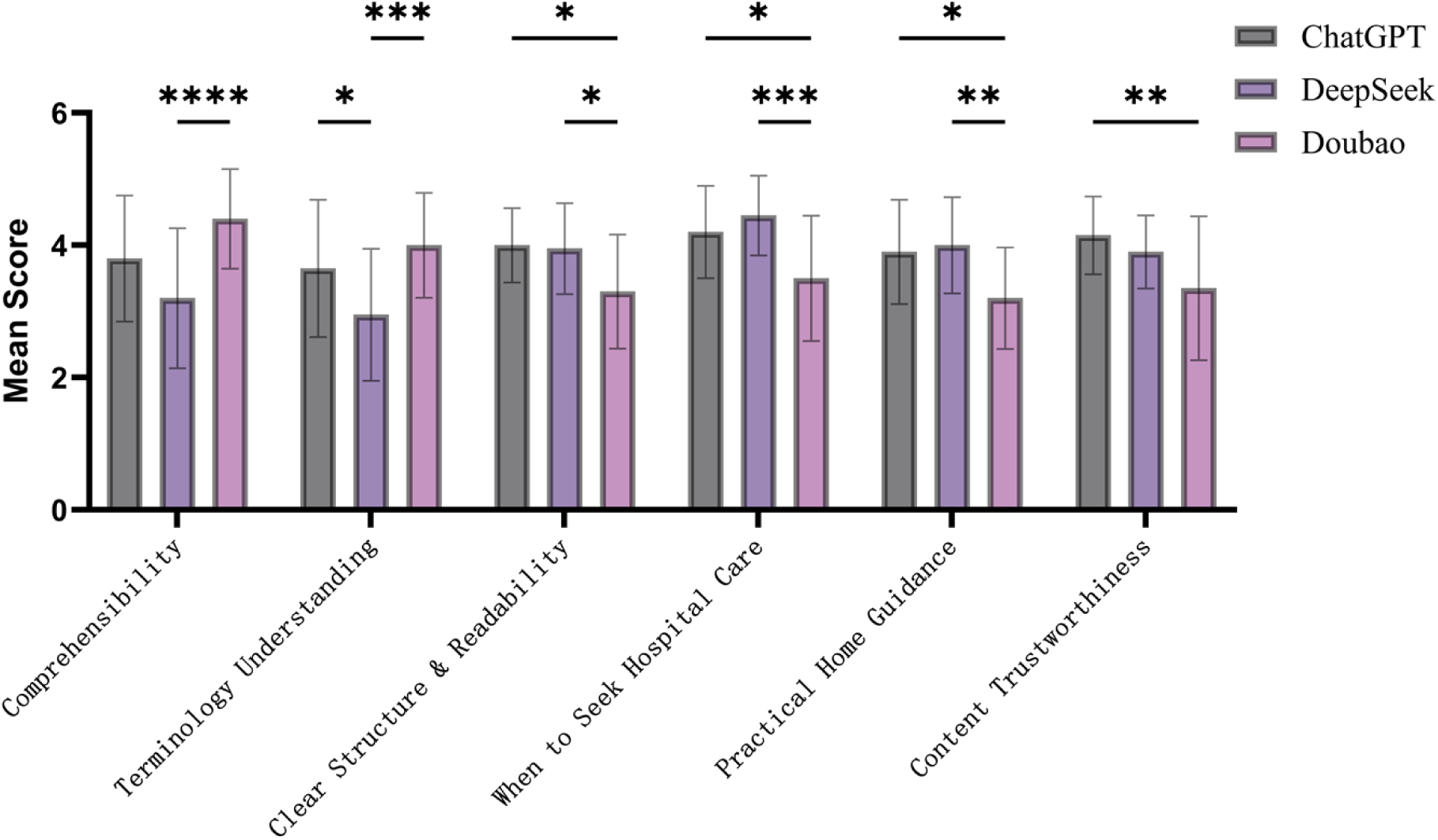
Intergroup comparison of patient survey scores across six dimensions. Bar charts presenting scores from 20 patients evaluating ChatGPT, DeepSeek, and Doubao across six dimensions (1–5 Likert scale). For “comprehensibility” and “terminology understanding,” Doubao scored significantly higher than ChatGPT and DeepSeek (****P < 0.0001, ***P < 0.001, *P < 0.05). For “clear structure,” “when to seek hospital care,” “practical home guidance,” and “content trustworthiness,” ChatGPT and DeepSeek did not differ significantly, while both scored significantly higher than Doubao (***P < 0.001, **P < 0.01, *P < 0.05).

**Figure 9.**
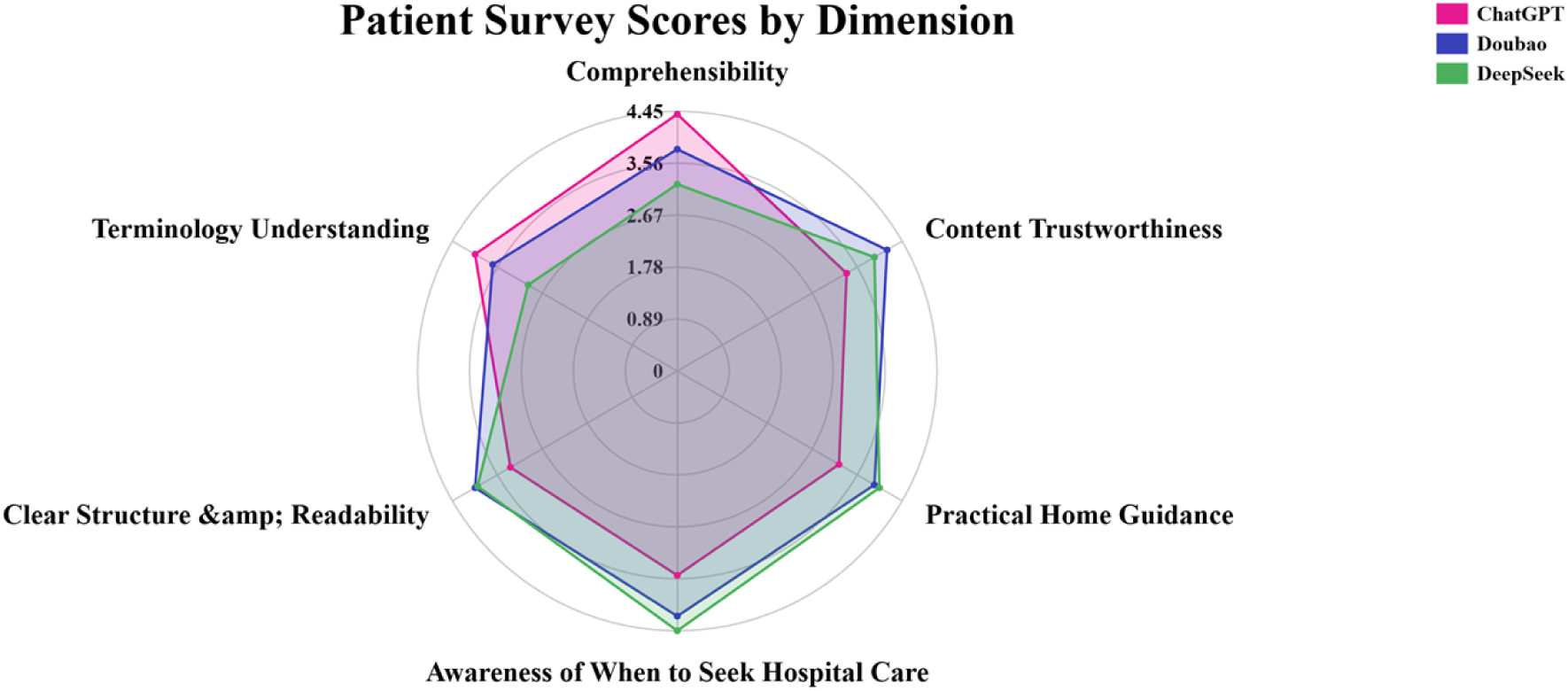
Radar chart of patient survey scores across six evaluation dimensions. Radar chart illustrating the overall performance of ChatGPT, DeepSeek, and Doubao across six dimensions: comprehensibility, terminology understanding, clear structure and readability, awareness of when to seek hospital care, practical home guidance, and content trustworthiness. ChatGPT and DeepSeek demonstrated superior performance in structural clarity, practical medical guidance, and content credibility. Doubao excelled in language plainness and terminology accessibility. DeepSeek exhibited the most balanced performance across all dimensions.

**Figure 10.**
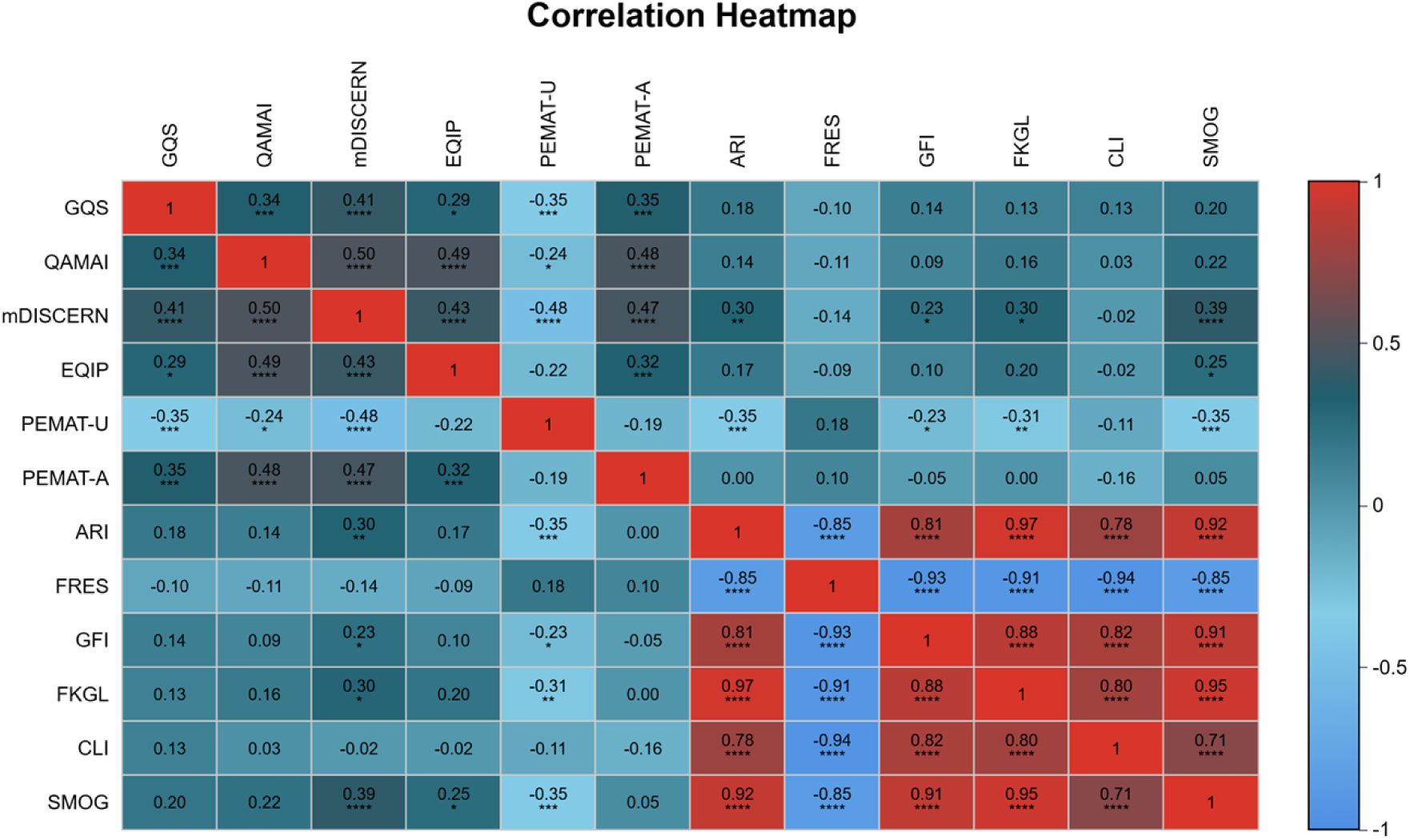
Spearman correlation heatmap of twelve evaluation metrics. Heatmap displaying Spearman correlation coefficients among GQS, QAMAI, mDISCERN, EQIP, PEMAT-U, PEMAT-A, and six readability indices (ARI, FRES, GFI, FKGL, CLI, SMOG). Strong positive correlations were observed among ARI, GFI, FKGL, CLI, and SMOG (r = 0.71–0.97, P < 0.001), while FRES showed strong negative correlations with these indices (r = −0.85 to −0.94, P < 0.001). PEMAT-U was negatively correlated with most quality metrics (r = −0.22 to −0.48). *P < 0.05, **P < 0.01, ***P < 0.001.

**Table 6.** Distribution and Proportion of Qualitative Deficiencies in Response Content.

| Content | N(%) |
| --- | --- |
| Output Error | 0% |
| Hallucination | 18.18% |
| Omissions | 36.36% |
| Outdated Medical Advice | 45.46% |

**Table 7.** Demographic Characteristics of Patient Survey Participants.

| Characteristic |  | *n* | % |
| --- | --- | --- | --- |
| Sex | Male | 11 | 55.0 |
|  | Female | 9 | 45.0 |
| Age group | 18–30 years | 5 | 25.0 |
|  | 31–45 years | 7 | 35.0 |
|  | 46–60 years | 5 | 25.0 |
|  | ≥60 years | 3 | 15.0 |
| Education level | Junior high school or below | 4 | 20.0 |
|  | Senior high school / vocational school | 4 | 20.0 |
|  | College / bachelor's degree | 9 | 45.0 |
|  | Master's degree or above | 3 | 15.0 |
| Medical background | Yes | 4 | 20.0 |
|  | No | 16 | 80.0 |

**Table 8.** Patient Subjective Evaluation Scores and Preferences.

| Dimension | ChatGPT | DeepSeek | Doubao |
| --- | --- | --- | --- |
| Comprehensibility (6 items) | $3.62 \pm 0.38$ | $3.74 \pm 0.28$ | $3.95 \pm 0.27$ |
| Credibility (1 item) | $3.35 \pm 1.09$ | $3.90 \pm 0.55$ | $4.15 \pm 0.59$ |
| Overall preference, n (%) | 3 (15.0) | 7 (35.0) | 9 (45.0) |

### 3.6 Patient Survey Results

A total of 20 patients with a personal or family history of bee stings were enrolled for subjective evaluation. The sample comprised 11 men (55.0%) and 9 women (45.0%). Age distribution: 18–30 years, 5 (25.0%); 31–45 years, 7 (35.0%); 46–60 years, 5 (25.0%); and over 60 years, 3 (15.0%). Education level: junior high school or below, 4 (20.0%); senior high school or vocational school, 4 (20.0%); college or bachelor’s degree, 9 (45.0%); and master’s degree or above, 3 (15.0%). Four patients (20.0%) had a medical background.

Regarding overall preference, 3 of the 20 patients (15.0%) preferred ChatGPT’s responses, 7 (35.0%) p referred DeepSeek, 9 (45.0%) preferred Doubao, and 1 (5.0%) expressed no preference or unwillingne ss to reference any of the responses. Preference analysis revealed that those choosing ChatGPT did so primarily because it ^“^told me exactly what to do^”^ and ^“^explained more clearly so I could understand.^”^ D eepSeek was preferred mainly because it ^“^felt more professional and credible.^”^ Doubao was selected fo r ^“^telling me exactly what to do,^”^ being ^“^shorter and less verbose,^”^ and providing ^“^more detailed and comprehensive content.^”^

### 3.7 Correlation Analysis Across Metrics

Spearman correlation analysis was performed on all assessment metrics, with results presented as a heatmap. Within the quality and reliability metrics, moderate to strong positive correlations were generally observed. The strongest correlation was between mDISCERN and QAMAI (r = 0.50, P < 0.0001); GQS also correlated significantly with mDISCERN (r = 0.41, P < 0.0001) and QAMAI (r = 0.34, P < 0.001). Notably, PEMAT-U was negatively correlated with most quality metrics, with a correlation coefficient of −0.48 with mDISCERN (P < 0.0001), suggesting a potential negative correlation between understandability and information reliability. Within the readability indices, ARI and FKGL showed a very strong positive correlation (r = 0.97, P < 0.0001), and FRES was strongly negatively correlated with the other readability indices (r = −0.85 to −0.94), consistent with expectations. Correlations between quality metrics and readability indices were generally weak, indicating that information quality and text readability are relatively independent dimensions.

## 4. Discussion

### 4.1 Principal Findings

This study evaluated the information quality, reliability, readability, and content deficits of three mainstream LLMs—ChatGPT, DeepSeek, and Doubao—in the domain of bee stings. Several key findings emerged. First, the three models showed distinct strengths across quality and reliability metrics: DeepSeek significantly outperformed the others on mDISCERN, ChatGPT led on QAMAI and EQIP, and both surpassed Doubao on most indicators. Second, Doubao paradoxically performed best on understandability (PEMAT-U), revealing an inverse relationship between quality and comprehensibility. Third, the readability levels of all models were significantly above recommended standards. Fourth, outdated medical advice was the most prevalent content deficit type, suggesting that LLMs have a lag in knowledge updating.

### 4.2 Quality and Reliability

DeepSeek achieved the highest mDISCERN score (median 21.00), significantly surpassing ChatGPT (18.00) and Doubao (13.00). The mDISCERN instrument primarily evaluates source reliability, information balance, and the acknowledgment of uncertainty; DeepSeek’s stronger performance in these areas may reflect targeted training on Chinese medical corpora and a greater tendency to cite evidence-based sources ^[27]^. ChatGPT, however, attained the highest QAMAI (median 25.00) and EQIP (75.00%) scores, indicating superior performance in overall information quality and patient information standards. This finding is consistent with research in allergology ^[28]^ and emergency medicine ^[30]^, which similarly reported better quality indicators for ChatGPT.

Doubao ranked lowest on all quality and reliability metrics, with a mDISCERN median of only 13.00 and an EQIP median of 62.50%, placing it at the lower boundary of the ^“^good quality, minor shortcomings^”^ classification. Although Doubao’s GQS median was 4.00, the gap relative to the 5.00 achieved by ChatGPT and DeepSeek suggests substantial room for improvement in information completeness and source transparency.

### 4.3 Understandability and Actionability

Notably, Doubao performed best on understandability (PEMAT-U median 73.00%), significantly outperforming DeepSeek (65.00%), with ChatGPT in between (71.00%). Although counterintuitive, this result may be attributable to Doubao’s response style—it tends to generate more concise, colloquial answers that, while less comprehensive, lower the barrier to comprehension. Similarly, Zhang et al. ^[25]^ found in their study of pediatric anesthesia that the most detailed model was not necessarily the most understandable.

For actionability, ChatGPT and DeepSeek performed comparably (PEMAT-A median 60.00%), both significantly outperforming Doubao (40.00%). PEMAT-A evaluates whether information can guide patients to take specific actions; Doubao’s weakness in this dimension may stem from its overly brief responses and lack of concrete action steps.

### 4.4 Readability

All models fell well short of recommended readability standards. The American Medical Association recommends that health materials intended for the public achieve a sixth- to seventh-grade reading level ^[29]^, yet the FKGL for all models in this study exceeded grade 10 and the SMOG exceeded grade 9. DeepSeek exhibited the greatest reading difficulty (FKGL = 12.48, ARI = 13.88), likely owing to its tendency to employ more specialized terminology and more complex sentence structures. Doubao had relatively better readability (SMOG = 9.46) but still fell far short of the recommended standard.

FRES, GFI, and CLI showed no statistically significant differences among the three groups, whereas ARI, FKGL, and SMOG differed significantly, suggesting that different readability formulae exhibit varying sensitivity to Chinese text. This finding indicates that multiple indices should be used in combination when evaluating the readability of Chinese health information, rather than relying on any single metric.

### 4.5 Content Deficits

Content deficit analysis revealed that outdated medical advice was the most common deficit type (45.45%), a finding of particular concern in the bee sting domain. Outdated recommendations included: no longer recommended methods for removing bee stingers (e.g., tweezers vs. scraping), outdated antihistamine recommendations, and outdated descriptions of the timing of epinephrine administration. These outdated suggestions may originate from older popular science articles and obsolete guidelines in the LLMs’ training data, reflecting a lag in knowledge updating.

Omissions of critical information accounted for 36.36%; the missing content primarily included: distinguishing features between local reactions and systemic anaphylaxis after a bee sting, specific steps for using an epinephrine auto-injector, and emergency management protocols for patients with a known allergy history who are re-stung. Hallucinations constituted 18.18%, mainly manifesting as fabricated drug dosages or fictitious statistics ^[31][32]^ The absence of output errors suggests that all three models possess a certain degree of reliability in basic medical facts.

### 4.6 Clinical Implications

The findings of this study yield the following implications for clinical practice and public health, and align with the LLM health information reporting standards proposed by Huo et al.^[30]^. Clinicians should be aware that patients may obtain bee sting information through LLMs and should proactively supplement content that AI may have omitted (e.g., timing of epinephrine administration, early recognition of anaphylaxis) and correct outdated recommendations (e.g., stinger removal methods). For public health authorities, this study highlights the readability barriers and knowledge-update lags in LLM-generated first-aid information intended for the public, suggesting a need for guidelines governing the use of LLMs in health information dissemination. For LLM developers, specialized training in the domain of anaphylaxis first aid is warranted, along with the establishment of knowledge-update mechanisms to reduce outdated medical advice and the development of tiered language generation strategies tailored to users with varying levels of health literacy.

## 5. Limitations

This study has several limitations. First, only 25 questions were evaluated, and the sample size may not comprehensively cover all clinical scenarios related to bee stings. Second, the assessments were conducted at a single time point, and LLMs may undergo temporal drift owing to platform updates. Third, differences in model versions and access methods may affect comparability. Fourth, human expert responses were not included as a reference standard. Fifth, content deficit classification relied on subjective expert judgment. Sixth, the patient survey had a small sample size (*n* = 20), limiting statistical power.

## 6. Conclusion

This study evaluated the information quality, reliability, readability, and content deficits of three mainstream LLMs—ChatGPT, DeepSeek, and Doubao—in the domain of bee stings. The results indicate that the three models exhibited distinct strengths in quality and reliability: DeepSeek achieved the highest DISCERN score, suggesting advantages in source reliability and information balance; ChatGPT led in QAMAI and EQIP, reflecting superior overall quality and patient information standards; and Doubao ranked lowest on all quality indicators but paradoxically performed best on understandability, revealing an inverse quality–understandability relationship. None of the models met the readability standards recommended for public-facing health information. Content deficit analysis identified outdated medical advice as the predominant deficit type (45.45%), indicating that current LLMs lack up-to-date knowledge on bee stings.

On the basis of these findings, clinicians should guide patients in the judicious use of LLMs for bee sting information, proactively supplement critical content, and correct outdated recommendations. LLM developers should pursue targeted optimization for the anaphylaxis first-aid domain, with emphasis on establishing knowledge-update mechanisms, enhancing information completeness, and developing adaptive readability strategies for users with varying levels of health literacy. Future research should expand the question set, include human clinical experts as a reference standard, and conduct patient surveys to evaluate the actual impact of LLM-generated information on patient behavior.

## Data Availability

The datasets used and analyzed during the current study are available from the corresponding author upon reasonable request.

https://readabilityformulas.com/readability-scoring-system.php

## 7. Ethical Statement

This study was approved by the Ethics Committee of People’s Hospital of Yuechi County (Approval N o. 2026-Ethical Approval-012-01). Informed consent was obtained from all participants via the first page of the questionnaire prior to participation. This survey collected only anonymous responses without any personally identifiable information. The study was conducted in accordance with the Declaration of Helsinki.

## 8. Acknowledgments

The authors would like to express their sincere gratitude to the editorial team and anonymous reviewers for their valuable feedback and insightful comments, which have significantly improved the quality of this manuscript.

## References

[1] Golden, D. B. K., & Carter, M. C. (2019). Insect Sting Anaphylaxis-Or Mastocytosis-Or Something Else?. The journal of allergy and clinical immunology. In practice, 7(4), 1117–1123. 10.1016/j.jaip.2019.01.014

[2] Bilò, M. B., & Bonifazi, F. (2009). The natural history and epidemiology of insect venom allergy: clinical implications. Clinical and experimental allergy : journal of the British Society for Allergy and Clinical Immunology, 39(10), 1467–1476. 10.1111/j.1365-2222.2009.03324.x

[3] Golden, D. B., Demain, J., Freeman, T., et al. (2017). Stinging insect hypersensitivity: A practice parameter update 2016. Annals of allergy, asthma & immunology : official publication of the American College of Allergy, Asthma, & Immunology, 118(1), 28–54. 10.1016/j.anai.2016.10.031

[4] Thirunavukarasu, A. J., Ting, D. S. J., Elangovan, K., et al. (2023). Large language models in medicine. Nature Medicine, 29(8), 1930–1940. 10.1038/s41591-023-02448-8

[5] Biswas, S. S. (2023). Role of Chat GPT in public health. Annals of Biomedical Engineering, 51(5), 868–869. 10.1007/s10439-023-03172-7

[6] Dede, B. T., Oğuz, M., Alyanak, B., et al. (2025). Competencies of large language models about piriformis syndrome: Quality, accuracy, completeness, and readability study. HSS Journal, 21(3), 342–347. 10.1177/15563316251340697

[7] Huang, J., Yu, H., Chen, J., et al. (2026). Can large language models be trusted? Reliability and readability of responses to perinatal depression FAQs. Frontiers in Public Health, 14, 1760872. 10.3389/fpubh.2026.1760872

[8] Lakdawala, N., Channa, L., Gronbeck, C., et al. (2023). Assessing the accuracy and comprehensiveness of ChatGPT in offering clinical guidance for atopic dermatitis and acne vulgaris. JMIR Dermatology, 6, e50409. 10.2196/50409

[9] Musheyev, D., Pan, A., Loeb, S., et al. (2024). How well do artificial intelligence chatbots respond to the top search queries about urological malignancies? European Urology, 85(1), 13–16. 10.1016/j.eururo.2023.07.004

[10] von Elm, E., Altman, D. G., Egger, M., et al. (2007). Strengthening the Reporting of Observational Studies in Epidemiology (STROBE) statement: Guidelines for reporting observational studies. BMJ, 335(7624), 806–808. 10.1136/bmj.39335.541782.AD

[11] CHART Collaborative. (2025). Reporting guidelines for chatbot health advice studies: Explanation and elaboration for the Chatbot Assessment Reporting Tool (CHART). BMJ, 390, e083305. 10.1136/bmj-2024-083305

[12] Chinese Society of Allergy. (2022). Chinese guidelines for the diagnosis and treatment of allergic reactions. National Medical Journal of China, 102(24), 1833–1848.

[13] Krishna, M. T., Ewan, P. W., Diwakar, L., et al. (2011). Diagnosis and management of hymenoptera venom allergy: British Society for Allergy and Clinical Immunology (BSACI) guidelines. Clinical & Experimental Allergy, 41(9), 1201–1220. 10.1111/j.1365-2222.2011.03788.x

[14] Bernard, A., Langille, M., Hughes, S., et al. (2007). A systematic review of patient inflammatory bowel disease information resources on the World Wide Web. The American Journal of Gastroenterology, 102(9), 2070–2077. 10.1111/j.1572-0241.2007.01325.x

[15] Charnock, D., Shepperd, S., Needham, G., & Gann, R. (1999). DISCERN: An instrument for judging the quality of written consumer health information on treatment choices. Journal of Epidemiology and Community Health, 53(2), 105–111. 10.1136/jech.53.2.105

[16] Robillard, J. M., Jun, J. H., Lai, J. A., et al. (2018). The QUEST for quality online health information: Validation of a short quantitative tool. BMC Medical Informatics and Decision Making, 18(1), Article 87. 10.1186/s12911-018-0668-9

[17] Moult, B., Franck, L. S., & Brady, H. (2004). Ensuring quality information for patients: Development and preliminary validation of a new instrument to improve the quality of written health care information. Health Expectations, 7(2), 165–175. 10.1111/j.1369-7625.2004.00273.x

[18] Shoemaker, S. J., Wolf, M. S., & Brach, C. (2014). Development of the Patient Education Materials Assessment Tool (PEMAT): A new measure of understandability and actionability for print and audiovisual patient information. Patient Education and Counseling, 96(3), 395–403. 10.1016/j.pec.2014.05.027

[19] Smith, E. A., & Kincaid, J. P. (1970). Derivation and validation of the automated readability index for use with technical materials. Human Factors, 12(5), 457–564. 10.1177/001872087001200505

[20] Flesch, R. (1948). A new readability yardstick. Journal of Applied Psychology, 32(3), 221–233. 10.1037/h0057532

[21] Gunning, R. (1952). The technique of clear writing. McGraw-Hill.

[22] Chinn, L., McGuirt, S., & Puri, S. (2014). The readability of online patient education materials from major anesthesiology associations and the American Society of Anesthesiologists. Open Journal of Anesthesiology, 4(1), 1–7. 10.4236/ojanes.2014.41001

[23] Coleman, M., & Liau, T. L. (1975). A computer readability formula designed for machine scoring. Journal of Applied Psychology, 60(2), 283–284. 10.1037/h0076540

[24] McLaughlin, G. H. (1969). SMOG grading: A new readability formula. Journal of Reading, 12(8), 639–646.

[25] Zhang, X., Dai, Y., Zhao, X., et al. (2026). Large language model chatbots as sources of pediatric anesthesia health advice: An evaluation of reliability and readability. Digital Health, 12, 20552076261464749. 10.1177/20552076261464749

[26] Koo, T. K., & Li, M. Y. (2016). A guideline of selecting and reporting intraclass correlation coefficients for reliability research. Journal of Chiropractic Medicine, 15(2), 155–163. 10.1016/j.jcm.2016.02.012

[27] Jiang, L., Chen, J., Lu, L., et al. (2025). Benchmarking Chinese medical LLMs: A MedBench-based analysis of performance gaps and hierarchical optimization strategies. arXiv. https://arxiv.org/abs/2503.07306

[28] Mathes, S., Seurig, S., Bluhme, F., et al. (2025). ChatGPT performance on 120 interdisciplinary allergology questions: Systematic evaluation with clinical error impact assessment. The Journal of Allergy and Clinical Immunology: In Practice. 10.1016/j.jaip.2025.03.030

[29] Weiss, B. D. (2007). Health literacy and patient safety: Help patients understand (2nd ed.). American Medical Association Foundation.

[30] Huo, B., Cai, T., & Xu, H. (2023). Reporting standards for the use of large language model-linked chatbots for health advice. Nature Medicine, 29(12), 2954–2955. 10.1038/s41591-023-02656-2

[31] Tazza, A., Sarker, A., & Pal, A. (2025). A framework to assess clinical safety and hallucination rates of LLMs for medical text summarisation. npj Digital Medicine, 8, Article 168. 10.1038/s41746-025-01670-7

[32] Chelli, M., Descamps, J., Claude, V., et al. (2024). Hallucination rates and reference accuracy of ChatGPT and Bard for systematic reviews. Journal of Medical Internet Research, 26, e53164. 10.2196/53164

